## Supplementary material for "CoViD-19, learning from the past: A wavelet and cross-correlation analysis of the epidemic dynamics looking to emergency calls and Twitter trends in Italian Lombardy region": Methods technical details

**Continuous wavelet transform**

The continuous wavelet transform (CWT) of a time function *x_t_* can be defined as the convolution of *x_t_* with a scaled and translated version of the wavelet function *ψ(t)* [39], that is equivalent to:

$X_{(a,b)}=\frac{1}{\sqrt{a}}\int_{-\infty}^{+\infty} x(t)\psi^{*}\left( \frac{t-b}{a} \right)dt$,	(1)

where:

- *b* is a translation parameter indicating locality in time;
- *a* is a scale parameter indicating locality in frequency;
- *t* is the dimensionless time;
- and the superscript ^*^ indicates the complex conjugate.

The CWT compares a signal with shifted and scaled (stretched or shrunk) copies of a basic wavelet. Scaling occurs in both the sense of contraction (shrunk) and of dilation (stretch). By changing the scale *a* and through a translation (parameter *b*) along the localised time index *t*, it is possible to represent the amplitude versus the scale and its variation with time [23]. The scale parameter is typically discretised to a fractional power of 2, whereas the translation parameter is discretised to integer values. To make different transforms and different time series comparable, it is necessary to normalize the wavelet function at each scale, thus obtaining unit energy [23].

Since we were interested in time series analysis with expected high variability and in phase information, a non-orthogonal complex transform was chosen and the following analytic Morlet (Gabor) wavelet was used:

$\psi\left( t \right)=\pi^{{-1}/4}e^{{-t^{2}}/2}e^{i\omega_{0}t}$,	(2)

where:

- *t* is dimensionless time;
- and *ω_0_* is the dimensionless wavelet central frequency.

In the analysis of a finite series sampled with a given sampling period (1 day in this specific instance), *t* is substituted by the time index *n*, *x(t)* is substituted with *x_n_* and the integral in (1) is substituted with a finite summation. Moreover, an appropriate normalization is introduced. One should also consider the so-called edge effects. Indeed, at the borders of the power spectrum, errors occur due to the finiteness of the time series: the cone of influence is the region of the spectrum where boundaries affect the values of the spectrum.

Here we have taken *ω_0_* = 6 to satisfy admissibility criteria (zero mean, locality in the time-scale plane). Morlet wavelet is a good choice for feature extraction since it provides a good balance between time and frequency localization [40-42], because – accordingly to Heisenberg uncertainty principle – time and frequency resolution are inversely proportional.

**Wavelet cross-spectrum**

The wavelet cross-spectrum (WCS) is a measure of the distribution of power of two signals. The WCS of the two time series, “daily infected” and the potential predictor one, is:

$C_{xy}\left( a,b \right)=S(C_{x}^{*}\left( a,b \right)C_{y}\left( a,b \right))$,	(3)

where:

- *x* stands for the daily infected time series;
- *y* indicates the predictor time series;
- *C_x_(a,b)* and *C_y_(a,b)* denote the CWTs of *x* and *y* at scales *a* and positions *b*;
- the superscript *^*^* is the complex conjugate;
- and *S* is a smoothing operator in time and scale.

**Wavelet coherence**

Magnitude-squared wavelet coherence (MSWC) is a measure of the correlation between two signals in the time-frequency plane. The MSWC of the two time series, “daily infected” and the predictor one, is:

${MSWC}_{xy}\left( a,b \right)=\frac{\left| S(C_{x}^{*}\left( a,b \right)C_{y}\left( a,b \right)) \right|^{2}}{S(\left| C_{x}\left( a,b \right) \right|^{2})\cdot S(\left| C_{y}\left( a,b \right) \right|^{2})}$,	(4)

where:

- *x* stands for the daily infected time series;
- *y* indicates the predictor time series;
- *C_x_(a,b)* and *C_y_(a,b)* denote the CWTs of *x* and *y* at scales *a* and positions *b*;
- the superscript *^*^* is the complex conjugate;
- and *S* is a smoothing operator in time and scale.

The coherence has been computed over logarithmic scales.

To perform this analysis (WCS and MSWC) we used the MatLab function “wcoherence”. In the computation of MSWC, we kept default MatLab settings using 12 steps (voices per octave) each for the logarithmically distributed frequency values. The number of octaves depends logarithmically (base 2) on the number of samples of the time series. The number of voices per octave indicates the number of intermediate scales to double (i.e. to increase the scale by an octave), thus for a larger value of voices per octave we have a finer discretization of the scale parameter, a greater number of scales, and consequently a lower frequency inferior limit of the wavelet spectrum. The fine sampling of scales in the CWT typically results in a high-fidelity signal analysis, through which one can localize transients in the signal.

**Time delay computation through WCT analysis**

The phase relationship of the WCS values has been used to identify the relative phase shift between the two signals, and thus – using the value of the period corresponding to each frequency of the WCS – the relative time lag between the input signals at each time:

$\Delta\varphi={arg(C_{xy}\left( a,b \right))}/\pi$	(5)

$T=f^{-1}$	(6)

$\Delta\varphi:2\pi= \Delta t :T \Longrightarrow\Delta t=\frac{\Delta\varphi\cdot T}{2\pi}$,	(7)

where:

- *T* is the period;
- *f* is the frequency;
- *Δφ* is the phase shift angle between the two signals, indicated in radians;
- 2*π* represents the full circle;
- and *Δt* is the time delay between the two signals.

**Cross-correlation and time lag interval estimates**

**The Fisher’s *z*-transform**

The Fisher’s logarithmic *z*-transformation applied to the cross-correlation sequence allows us to obtain a sampling distribution of the resulting variable which is approximately normal:

$z_{j}=\frac{1}{2}\ln\left( \frac{1+r_{j}}{1-r_{j}} \right)=arctanh(r_{j})$,	(8)

where:

- *j* is the index of the time lag of the cross-correlation sequence;
- *z* is the transformed variable at the *j^th^* time lag;
- *r* is the cross-correlation value at the *j^th^* time lag.

Moreover, the corresponding variance is constant and stable over different values of the underlying true correlation. Indeed, the standard error of the transformed variable is approximately:

$s= \frac{1}{\sqrt{n-3}}$,	(9)

where:

- *s* is the standard error;
- *n* is the sample size.

It is clearly evident that *s* is independent of the value of the correlation, depending only on the sample size *n*.

The normalizing transformation of the distribution and the variance stabilizing effect are the main features that make this procedure suitable even for small sample sizes, as in the case of emergency calls time series [32].

The Fisher’s transformation and its exponential inverse thus can be used to construct confidence intervals for the cross-correlation sequence estimate using standard normal theory [32]. Particularly, confidence intervals can be calculated in *z* coordinates and the result can be reversely transformed as it follows:

$r_{j}=\frac{e^{2z_{j}}-1}{e^{2z_{j}}+1}=tanh(z_{j})$,	(10)

where:

- *j* is the index of the time lag of the cross-correlation sequence;
- *r* is the cross-correlation value at the *j^th^* time lag;
- *z* is the transformed variable at the *j^th^* time lag.

**Monte Carlo simulation through Fourier transform random phase test**

For the present Monte Carlo method, proposed by Ebisuzaki (1997) [33], we used a uniformly distributed random number generator. The aim was to build a model of the potential predictor with the same basic properties of the original signal. First of all, since MatLab algorithm uses a Discrete Fourier Transform (DFT) to compute the cross-correlation function in the frequency domain, we calculated the Fast Fourier Transform (FFT) of the predictor time series. Then, we generated random numbers representing the values of the phase of a signal: the number of random phases of each series was the same as the length of the original time sequence. After having created random phase sequences, we generated complex numbers through the following product:

$FFT\left( x_{k}^{*} \right)=|FFT\left( x_{k} \right)|e^{i\theta_{k}}$,	(11)

where:

- *k* is the frequency index;
- *FFT(x_k_^*^)* is the FFT of the surrogate time series;
- *FFT(x_k_)* is the FFT of the original time series;
- *θ_k_* is the randomly generated phase.

In this way we obtained a FFT sequence displaying the same power spectrum as the predictor time series [33]. This operation can be considered a sort of sampling in the frequency domain, differently from the bootstrap techniques which resample in the time domain [33]. Subsequently, we performed on this FFT sequence the Inverse Fast Fourier Transform (IFFT) so that we created the surrogate series converting frequency domain data into time domain samples. If we consider the algorithm used to compute the cross-correlation function, we have just built a time series with the same auto-correlation as the potential predictor. After that, we performed 1,000 simulations cross-correlating the surrogate series with the daily infected one. Finally, we computed the mean of the simulated cross-correlation sequences, the time delay corresponding to the peak of the simulated cross-correlation sequences mean, and their relative confidence intervals. This procedure, as already described, preserved the same properties of the original time series, such as power spectrum and auto-correlation [33].
